# Study protocol: HiSNAP trial – a multi-centre, randomised, open label, blinded end-point, safety and efficacy trial of conventional (300mg/kg) versus higher doses of acetylcysteine (450mg/kg and 600mg/kg) in patients with paracetamol overdose in the United Kingdom

**DOI:** 10.1101/2024.11.12.24317178

**Authors:** Christopher Humphries, Ellise Clarke, Michael Eddleston, Marianne Gillings, Sîan Irvine, Liza Keating, Anna Miell, Lynsey Milne, Laura Muir, Rachel O’Brien, Katherine Oatey, Rajendra Raman, Ruben Thanacoody, Sharon Tuck, Christopher J Weir, David M Wood, James W Dear, HiSNAP Trial Investigators

## Abstract

**Introduction:** In overdose, a larger proportion of paracetamol (acetaminophen) is converted in the liver to the toxic metabolite N-acetyl-p-benzoquinone imine (NAPQI). Glutathione (GSH) is the endogenous antioxidant that protects cells from NAPQI-induced injury. In overdose, GSH stores may become depleted, leaving NAPQI free to produce liver damage. Acetylcysteine (NAC) helps prevent paracetamol toxicity by replenishing liver GSH. This protective effect of NAC produces specific metabolites in the circulation. Currently, regardless of the paracetamol dose ingested, patients in the United Kingdom receive a dose of NAC based only on their weight. Basic pharmacology, mathematical modelling and observational studies suggests that this dose may be insufficient in some patients (particularly those taking a large overdose).

**Methods and analysis:** A multi-centre trial, taking place across several hospitals in Scotland, UK, within Emergency Departments and Acute Medical Units. Recruitment commenced 19 Feb 2024, and is anticipated to run for approximately two years. This is a three-group dose finding trial, in which participants are assigned in a 1:1:1 ratio to either standard NAC (300mg/kg), or higher doses of 450mg/kg (Group 1) and 600mg/kg (Group 2). The primary outcome is the proportion of paracetamol metabolites in the circulation that are directly produced by GSH/NAC detoxification of NAPQI. A higher proportion of these metabolites will indicate that the additional NAC is reducing the amount of toxic paracetamol metabolites in the body. The study will first test the primary outcome on the HiSNAP Group 2 against Standard NAC; only if that is significant, will HiSNAP Group 1 be tested against Standard NAC.

**Ethics and dissemination:** The HiSNAP trial has been approved by East Midlands (Derby) Research Ethics Committee (reference 23/EM/0129), NHS Lothian Research and Development department, and the MHRA. Results will be disseminated by peer-reviewed publication, conferences, and linked on isrctn.com.

**Registration details:** ISRCTN 17516192

**Strengths and limitations of this study:**

- The study will systematically collect data to assess the safety and efficacy of higher acetylcysteine doses and the current standard dose.
- The study primary endpoint is measurable in every patient, in contrast with paracetamol overdose trials examining liver injury (which only occurs in approximately 10% of patients).
- The study has pragmatic inclusion criteria – the decision that the patient requires acetylcysteine lies with the clinical team.
- The study seeks to minimise imbalances between treatment groups by stratifying randomisation according to whether a patient is at high risk of liver injury after overdose.
- The study is limited by the primary outcome being biomarker-based. Any benefits on clinical outcomes will still require confirmation.

## Introduction

### Background

Paracetamol (acetaminophen, APAP) overdose is a common medical emergency. Across the UK, approximately 100,000 people attend hospital following a paracetamol (acetaminophen, ‘APAP’) overdose every year (one every 5 minutes, same as myocardial infarction) and around half need admission for treatment with the antidote acetylcysteine (N-acetylcysteine – NAC).(1) Around 10% of these patients develop liver injury and approximately three people die from paracetamol induced liver failure every week.(2) Paracetamol overdose is more common in Scotland than any other UK region, or indeed globally.

### Rationale for study

There is a lack of robust evidence to guide treatment decisions. The Cochrane Library published a systematic review of interventions for paracetamol overdose in 2018.(3) The conclusion begins by saying “These results highlight the paucity of randomised clinical trials comparing different interventions for paracetamol overdose and their routes of administration and the low or very low level quality of the evidence that is available.” This situation has resulted in differences in treatment protocols that are not evidence-based and do not have adequate safety assessments. High dose NAC and repurposed, expensive, drugs are being recommended for off-label use, yet no randomised controlled trials (RCTs) have been performed for these treatment recommendations.(4)

Paracetamol is converted in the liver to the toxic metabolite N-acetyl-p-benzoquinone imine (NAPQI – Figure 1). Glutathione (GSH) is the endogenous antioxidant that protects cells from NAPQI-induced injury. In overdose, GSH stores are depleted leaving NAPQI free to produce liver damage. NAC prevents paracetamol toxicity by replenishing liver GSH. This protective effect of NAC produces specific metabolites in the circulation, shown in Figure 1. Currently, patients in the United Kingdom receive a dose of NAC that is based only on their weight. It is clear from the basic pharmacology, mathematical modelling and observational studies that some patients (particularly those taking a large overdose) may not be receiving enough NAC.(5-8)

**Figure 1.**
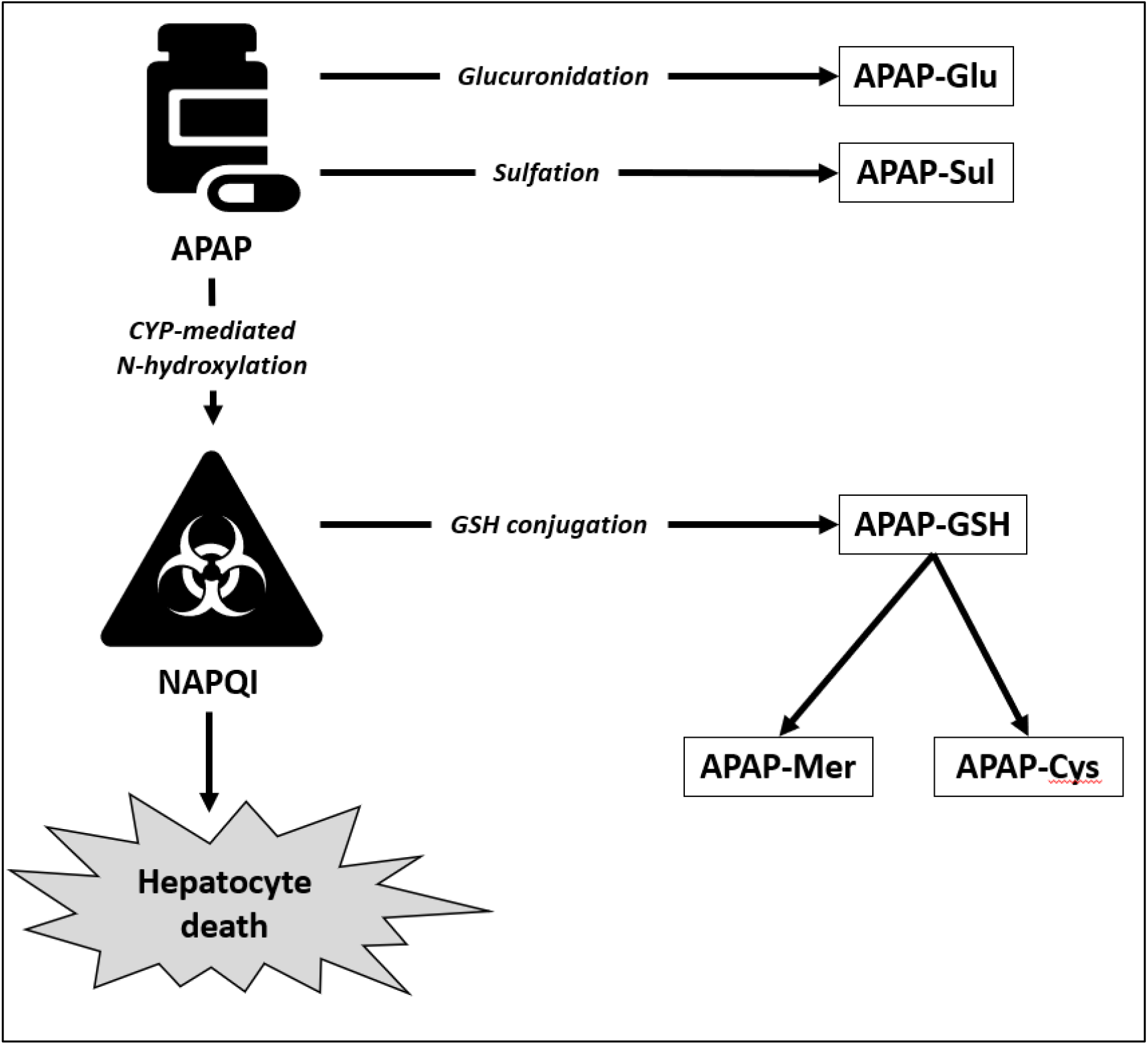
APAP (acetaminophen, paracetamol) metabolites. The study team have validated assays for APAP, APAP-Glu, APAP-Sul, APAP-GSH, APAP-Mer, and APAP-Cys.(9) In the 1970s a 20.25hr regimen for delivering NAC was developed in Edinburgh and elsewhere that became the worldwide standard-of-care.(10) The dose administered to patients using this regimen is limited by side-effects that are directly related to the high initial rate of the NAC intravenous infusion. We designed, trialled and implemented a new regimen for NAC (the ‘SNAP’ regimen) that delivers the same total dose as the 21hr regimen (300mg/kg) but with fewer dose-limiting side effects while being quicker to complete (12hr instead of 21hr).(11-13) This reduction in side effects means we can now safely test the effectiveness of higher doses. The SNAP regimen is endorsed by the UK National Poisons Information Service (NPIS) and the UK Royal College of Emergency Medicine and is now standard care across the UK and is used worldwide.(14) Some patients treated with the SNAP regimen require an additional infusion after 12h and receive a higher dose of 500 mg/kg over 22 hours. It is therefore timely to investigate the effectiveness of higher doses of NAC as current evidence is inconclusive.(13)

### Objectives

In this trial we will test the hypothesis that a higher NAC dose will increase detoxification of paracetamol. There are observational studies that suggest higher doses of NAC has potential to reduce liver injury in patients taking large overdoses, but no RCTs have been performed and safety has not been rigorously assessed.(9)

### Trial design

To assess the risk/benefit profile of high dose NAC, we propose a three group dose-finding trial as shown in Figure 2. The maximum dose to be studied is 600mg/kg, chosen because mathematical models predict this will be a sufficient dose to effectively treat 98% of patients.(6) The other treatment groups will receive 450mg/kg (1.5 x standard care) or 300mg/kg (standard care). The primary outcome is the proportion of paracetamol metabolites in the circulation that are directly produced by GSH/NAC detoxification of NAPQI (i.e. serum %GSH metabolite concentration after 12 hours of NAC). A higher proportion of these metabolites will indicate that the additional NAC is reducing the amount of toxic paracetamol metabolites in the body. For secondary endpoints, standard and novel liver injury markers will be measured to explore the effect of higher NAC doses on preventing liver injury. Safety will be assessed by systematically recording adverse events (AEs).

**Figure 2.**
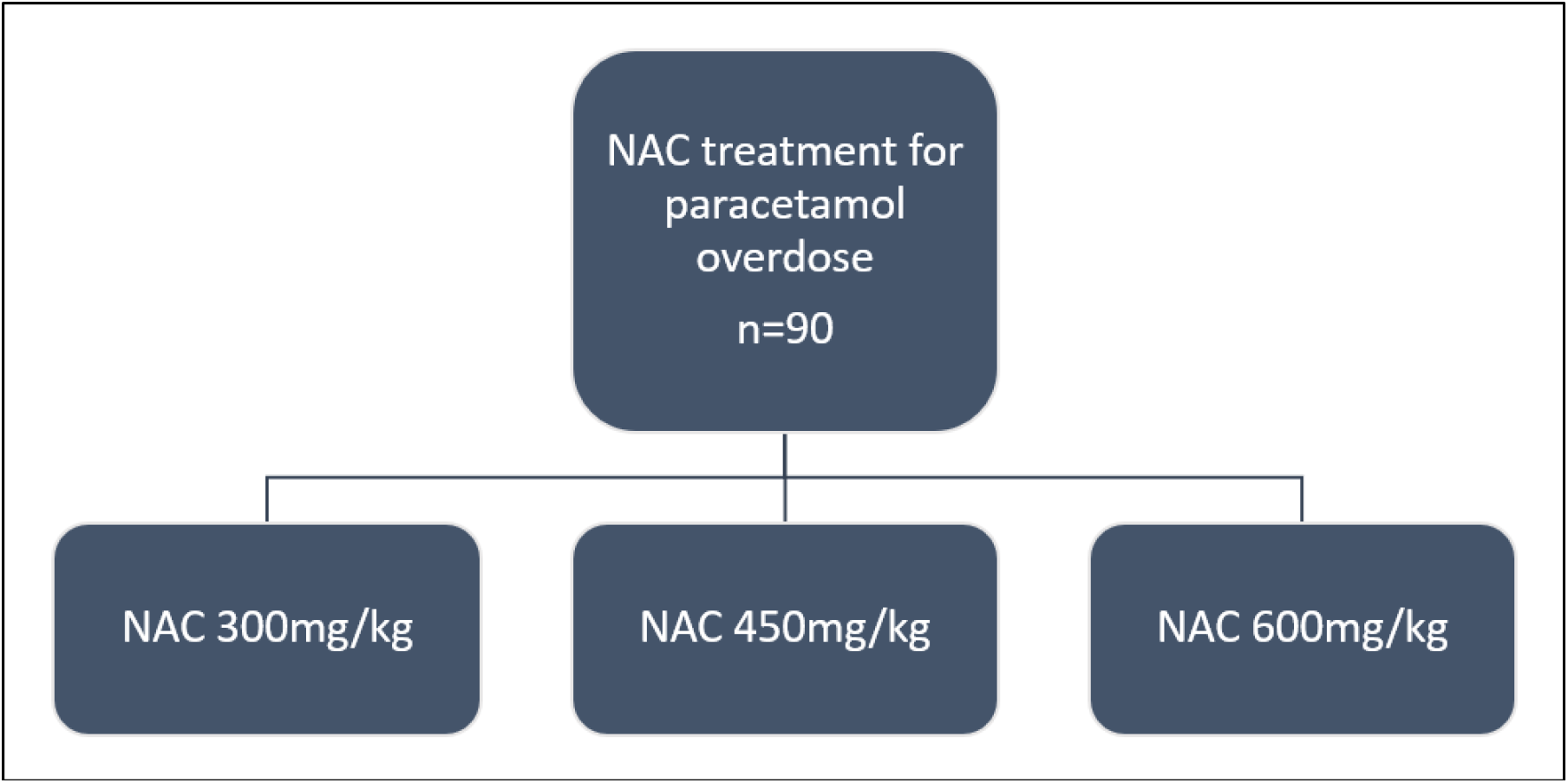
Trial design flow chart. Participants are randomised in a 1:1:1 ratio, with the potential to extend the sample size by up to 20% (i.e. maximum of 108 patients) if agreed by Trial Steering Committee, Sponsor, and funder.

## Methods

### Study setting

This is a multi-centre trial, taking place across several hospitals in Scotland, UK, within Emergency Departments and Acute Medical Units. Recruitment commenced 19 Feb 2024, and is anticipated to run for approximately two years. Eligible patients who have provided written informed consent will receive 12 hours of study intervention and will be followed up for seven days. Please refer to Figure 3 for a study visits flow diagram.

**Figure 3.**
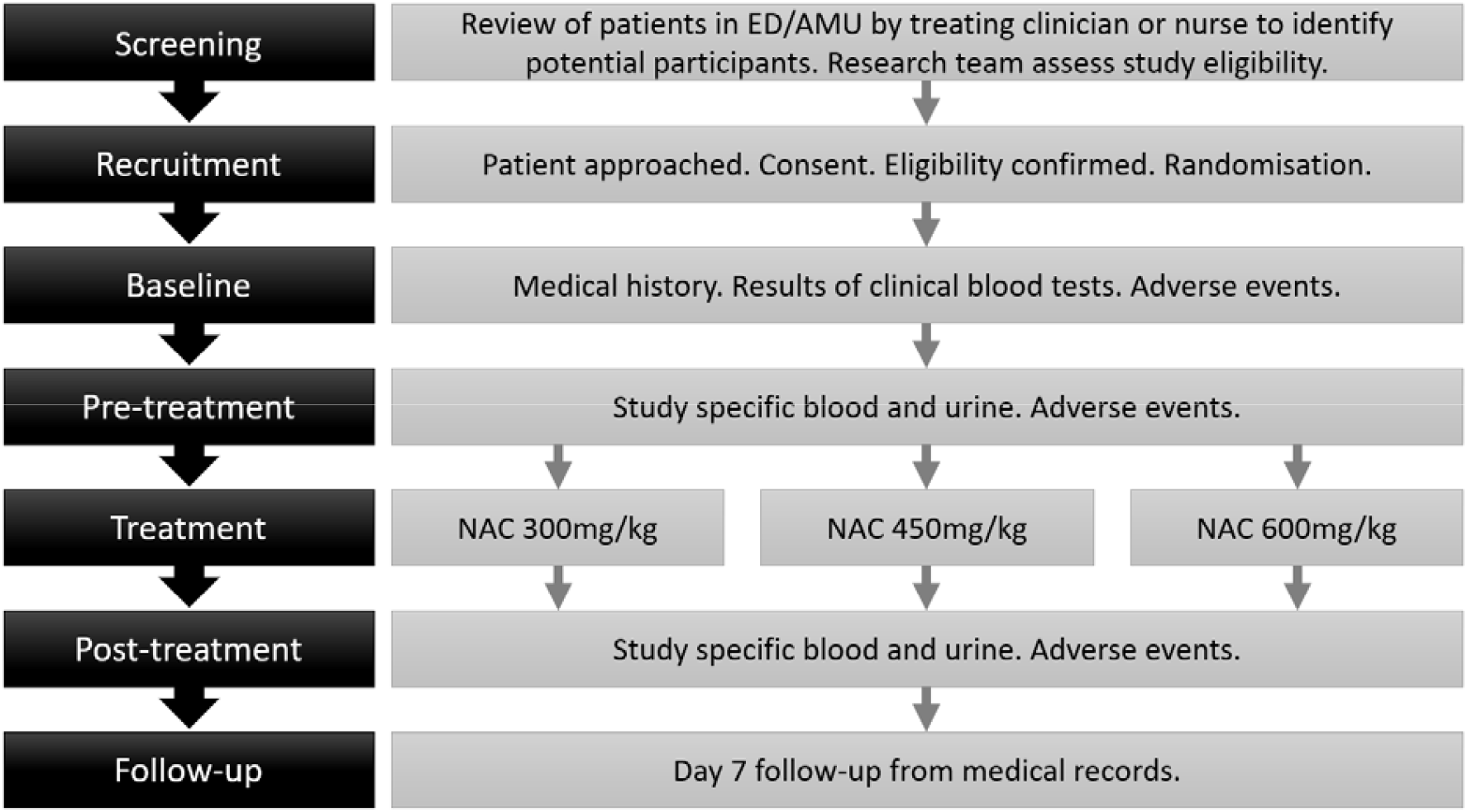
Study visits flow diagram

### Patient and public involvement

A Patient and Public Involvement group for Emergency Medicine Research (Sheffield, UK) reviewed and gave feedback on the study design which was actioned prior to grant application submission. In addition, a survey of patients presenting with acute overdose was performed to explore willingness to participate in clinical trials, which demonstrated no difference between this group and general medical patients.

### Eligibility criteria

#### Inclusion criteria

All of:

1. Paracetamol overdose presenting to hospital within 24hrs of taking their last dose of paracetamol. All patterns of overdose (single, staggered overdoses and therapeutic excess); accidental or deliberate; paracetamol alone or mixtures of tablets are eligible.
2. Patient deemed to need NAC treatment by the clinical team.
3. Blood paracetamol concentration and alanine aminotransferase (ALT) results are available (clinical care bloods).
4. Provision of informed consent.
5. Adult (16 years old or above).

#### Exclusion criteria

Any of:

1. Patients that do not have the capacity to consent.
2. Patients who are pregnant or breast feeding.
3. Patients who have previously participated in the study.
4. Patients who, in the opinion of the responsible clinician/nurse, are unlikely to complete the full course of NAC e.g. expressing wish to self-discharge.
5. Patients detained under the Mental Health Act.
6. Patients already started on NAC treatment.
7. Patients with known viral hepatitis or HIV.
8. Prisoners

### Interventions

The product under investigation is Acetylcysteine 200mg/ml Concentrate for Solution for Infusion, delivered by peripheral venous infusion per the SNAP regimen for the treatment of paracetamol poisoning. There will be three dosing groups:

1. Standard NAC as per the SNAP regimen (300mg/kg): initial loading dose (100mg/kg in 200mL) given intravenously over 2hr, followed by a second dose (200mg/kg in 1000 mL) infused over 10hr, OR
2. HiSNAP Group 1 (450mg/kg): initial loading dose (150mg/kg in 200 mL) given intravenously over 2hr, followed by a second dose (300mg/kg in 1000 mL) infused over 10hr, OR
3. HiSNAP Group 2 (600mg/kg): initial loading dose (200mg/kg in 200 mL) given intravenously over 2hr, followed by a second dose (400mg/kg in 1000 mL) infused over 10hr.

We will use the SNAP protocol for delivering NAC rather than the 21 hour protocol (as in the NAC marketing authorization) because the SNAP regimen is the UK standard of care endorsed by the Royal College of Emergency Medicine and the UK National Poisons Information Service.(14) All hospital regions in Scotland use the SNAP regimen to deliver NAC treatment. The only variation to standard practice is that NAC doses will be calculated for each patient using their body weight to the nearest kilogram (kg) rather than a 10kg weight-band as is currently used in clinical practice when patients receive standard NAC (300mg/kg).

There is no placebo arm.

An H1 antihistamine (Chlorphenamine 10 mg IV) and nebulised salbutamol may be administered to treat anaphylactoid reactions to NAC. Anti-emetic medicines may be given to treat nausea secondary to NAC. Any medication (prescription as well as over the counter drugs) or therapeutic intervention deemed necessary for the patient, and which, in the opinion of the Investigator, do not interfere with the safety and efficacy evaluations, may be continued.

### Outcomes

#### Primary outcome

The primary outcome measure of efficacy is the proportion of paracetamol metabolites in the circulation that are directly produced by GSH/NAC detoxification of NAPQI (i.e. (APAPCys+APAPMer+APAPGSH)/(APAPCys+APAPMer+APAPGSH+APAPGlu+APAPSul+APAP). A higher proportion of these metabolites will indicate that the additional NAC is reducing the amount of toxic paracetamol metabolites in the body.

#### Secondary outcomes

A full paracetamol metabolite panel (APAP-Cys, APAP-Glu, APAP-Sul, APAP-GSH, APAP-Mer) in serum and urine measured at baseline for serum and urine, 12 hours after the start of NAC for serum, and in the total urine collected in the 12 hour trial period will be conducted. Standard (ALT) and novel liver injury markers (keratin-18, miR-122) will be measured to explore the effect of higher NAC doses on preventing liver damage.(15) International Normalised Ratio (INR) will be measured as a marker of liver synthetic function.

Repeat hospital admission within seven days, acute liver failure, transfer to the Scottish Liver Transplantation Unit, liver transplantation and death will be reported.

To determine whether there are more adverse events with higher NAC doses, adverse reactions will be measured by a Likert scale on the following nine symptoms: nausea, feeling flushed, itchy skin, skin rash, chest pain, headache, feeling breathless, feeling wheezy, tongue/lip swelling. A free-text box is also available. This self-assessment questionnaire will be completed as a at baseline, and again 12 hours after starting NAC. The number of events and severity of reactions will be used to compare each dosing group.

Safety will be assessed by systematically recording adverse events (see ‘Safety assessments’). Anaphylactoid reactions and cerebral oedema are adverse events of special interest. The symptoms questionnaire will be reviewed as soon as possible after completion and within 7 days of infusion if the questionnaire is available. Any clinically significant events identified by an appropriately delegated physician will be recorded/reported as AEs/serious AEs (SAEs). A clinically significant event is defined as one which requires treatment and/or is recorded in the medical records.

### Recruitment

#### Identification

Patient screening will be undertaken by the nurses of the Emergency Medicine Research Group of Edinburgh (EMERGE). Participant eligibility will be verified by a delegated clinical trial physician after written informed consent has been obtained. Confirmation of eligibility will be recorded within the participants’ medical records and the participant added to the screening log.

The need for NAC treatment will be determined by the blood paracetamol concentration and liver function tests (including ALT) which are measured as part of routine care. Results from routine blood samples taken between 4 hours and 24 hours after last ingestion of paracetamol overdose may be used for this eligibility assessment.

Pregnancy testing will be carried out prior to randomisation if a potential participant is of child bearing potential and could be pregnant, and this will be recorded in the patient’s medical notes and the eCRF. Childbearing potential is defined as fertile, following menarche and until becoming postmenopausal unless permanently sterile. Permanent sterilisation methods include hysterectomy, bilateral salpingectomy and bilateral oophorectomy. A postmenopausal state is defined as no menses without an alternative medical cause for 12 months for women over 50 years old, and for 24 months for women under 50 years old. A high follicle stimulating hormone (FSH) level in the postmenopausal range may be used to confirm a post-menopausal state in women not using hormonal contraception or hormonal replacement therapy. However, in the absence of 12 months of amenorrhea, a single FSH measurement is insufficient.

#### Randomisation

Randomisation will be carried out using a web-based randomisation service (managed by the Edinburgh Clinical Trials Unit (ECTU)). Participants will be randomised by any appropriately delegated member of the trial team to Standard NAC (300mg/kg), HiSNAP Group 1 (450mg/kg) or HiSNAP Group 2 (600mg/kg) in a 1:1:1 ratio and minimised according to whether a patient meets the criteria of being at high risk of liver injury after overdose. High risk is defined as a hospital presentation ALT activity greater than upper limit of normal (defined as >50U/L) or a blood paracetamol concentration over the 300 line on nomogram (applies only to single acute overdoses - Figure 4). A random element will be included in the minimisation to maintain unpredictability.

**Figure 4.**
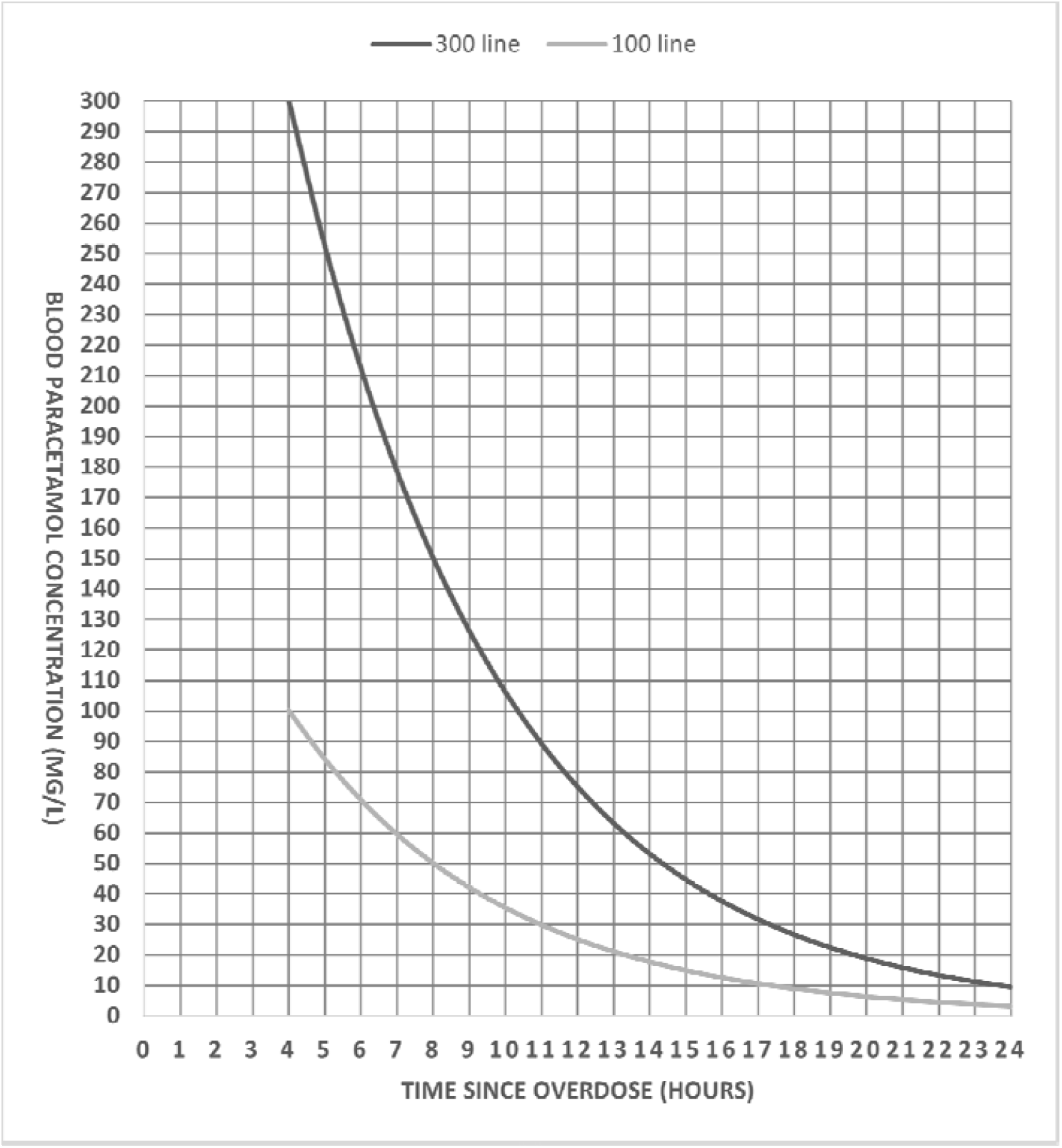
Nomogram used in UK clinical practice to determine if treatment is required for paracetamol overdose (100 line) and line used for trial purposes to determine if single acute overdoses are over the 300 line. Once a patient is randomised, they will remain in the study and have all outcomes recorded regardless of compliance with study allocation, unless they specifically withdraw consent to have data stored.

#### Blinding

Not applicable - the study is not blinded. The trial team members responsible for measuring the primary endpoint by mass spectrometry will be blinded to the treatment group of each sample.

#### Participant retention

As participation in the study does not extend significantly beyond the normal duration of treatment, and seven day follow-up is conducted using medical records, we do not anticipate significant issues with participant retention.

### Study assessments

### Use of study data

#### Data collection

Data will be collected from consent until 7 days after randomisation. Data items to be collected and data collection points are described in Table 1. The local research team as delegated will be responsible for data collection and recording this in the electronic case report form. Data will be collected by trained and delegated members of the research team.

**Table 1.**
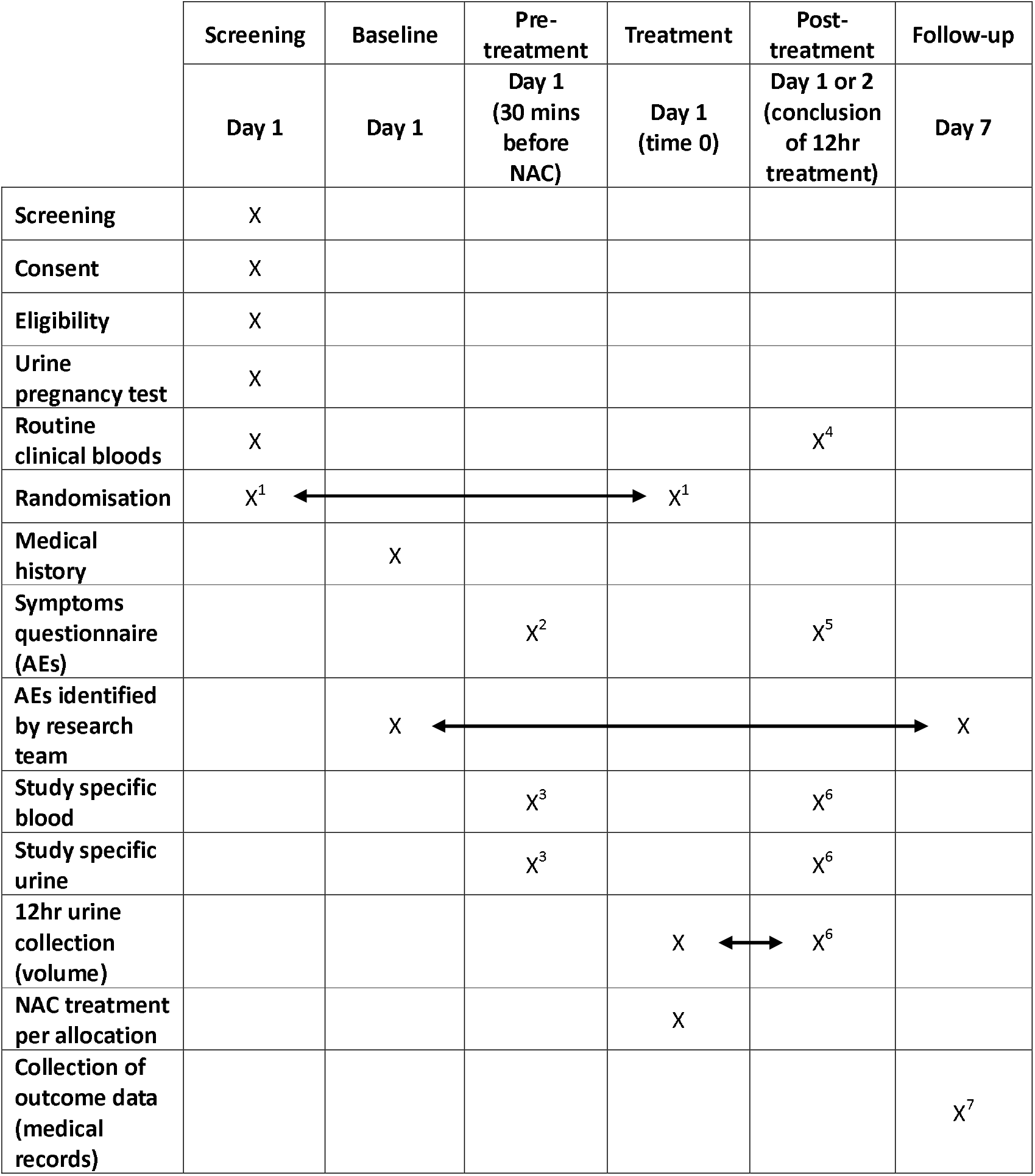
Study assessments. AE = Adverse Events. NAC = acetylcysteine. 1. Randomisation may take place at any time after consent but before NAC treatment depending on availability of blood results. 2. Baseline symptoms questionnaire 0-30mins before NAC treatment begins. 3. Pre-treatment interventions 30 minutes (+/- 15 min). 4. Routine bloods (+/- 1hr) routine clinical bloods - if bloods (or individual parameters) are not requested by the clinical team, this will not be recorded as a deviation. If a routine sample is not available, the research team may take an additional sample to measure the endpoint parameters. 5. 5.12hr Symptoms Questionnaire (+/- 2hr). Should ideally be collected as soon as possible after NAC treatment completed. If participant is discharged before questionnaire completed, the research team will make no more than three attempts to phone the patient and request the information retrospectively. 6. 12hr study specific samples (+/- 2hr). Should ideally be collected as soon as possible after 12hr NAC treatment completed. If participant refuses blood or urine sample collection this will not be recorded as a deviation but will be captured in eCRF. Urine: the participant will be asked to empty bladder at 12hr time point so final volume can be recorded on eCRF and research sample collected. Blood: where a study specific blood sample cannot be obtained, the research team will attempt to collect a sample from surplus clinical blood. 7. 7 Day follow up (+ 7 days). As day 7 follow up is collected from the medical records it can be reviewed and recorded in the eCRF up to 7 days after the time point, so it captures all relevant information.

#### Data management

Data collected about the participants will be entered into a password-protected, secure database (REDCap; http://www.project-redcap.org) provided and maintained by ECTU.(16,17) Trained and delegated members of the study team will be given password-protected logins to the database to complete data entry. No directly identifiable information will be entered into the electronic trial database.

Authorised personnel will be given protected access to relevant data. Consent is given from the participant. Examples of this include the Study Statistician in order to produce progress reports to the independent Data Monitoring Committee (DMC) and/or the interim or final statistical analyses; and the Trial Manager to produce blinded logistics reports; the Data Management Team to check the quality of the data, and the QA manager for audit and ongoing quality assurance reports); as well as representatives of the Sponsor for purposes of audit and monitoring or the regulatory authorities.

#### Data monitoring

The DMC will meet at least once each year, or once ten patients have been recruited into each trial arm, and subsequently when 20 patients have been recruited to each arm. The committee is an independent, multidisciplinary group with experience in the management of patients with paracetamol overdose, anticipated AEs, and the conduct and monitoring of randomised clinical trials. Additional urgent meetings may be arranged if any cases of cerebral oedema occur.

### Sample size and analysis plan

We have profiled paracetamol metabolites using samples from the SNAP and MAPP Trials (%GSH - mean 2.16% (SD 1.81)).(9) External validity is provided by a recent Australian cohort study that reported similar mean and SD for these metabolites.(18) To detect a 45% difference in this toxicokinetic marker between standard and each of the HiSNAP groups (cf 50% effect size estimated from mouse pilot data), 30 patients per group will give a power of 88% (log-transformation of data, 5% two-sided significance level, analysis by analysis of covariance adjusting for baseline value, assuming correlation 0.62 between baseline and post-NAC %GSH). To control the overall type I error at 5%, we will incorporate a hierarchical hypothesis testing structure. We will first test the primary outcome on the HiSNAP Group 2 against Standard NAC; only if that is significant, will we test HiSNAP Group 1 against Standard NAC. Patients with liver injury (and by definition excessive NAPQI production) have an increase in metabolites of between double to 4-fold (200-400%). Therefore, our study is powered to detect a clinically relevant effect size (45%).

The log-transformed primary outcome, %glutathione (GSH) metabolites, will be analysed using a normal linear model, adjusting for the pre-NAC %GSH value and for the continuous form of the baseline variables ALT and blood paracetamol concentration on which the minimisation factor is based. The treatment effect for each HiSNAP group versus Standard NAC will be estimated using the least squares mean difference and its 95% confidence interval (CI), back-transformed to the original scale to give the ratio for HiSNAP:Standard NAC.

Continuous secondary outcomes will be analysed in the same way as the primary outcome. Categorical secondary outcomes will be analysed using logistic regression, adjusting for the minimisation variables in the same way as in the normal linear model and reporting the treatment effect in each HiSNAP group versus Standard NAC by odds ratio (95% CI). A detailed statistical analysis plan will be finalised prior to database lock and unblinding of the trial statistician.

Exploratory pre-defined subgroup analyses will investigate the relationship between the treatment effect of each HiSNAP group and baseline levels of ALT and blood paracetamol level. Patients with a blood paracetamol concentration over 300 line on nomogram are at high risk of liver injury even with NAC treatment.(7) This will be done separately for each baseline variable by including an interaction term with treatment group in the normal linear model for the primary outcome.

### Safety assessments

Participants will be asked about the occurrence of AEs/SAEs at every visit during the study. Open ended and non-leading verbal questioning of the participant will be used to enquire about AE/SAE occurrence. Participants will also be asked if they have been admitted to hospital, had any accidents, used any new medicines or changed concomitant medication regimens. If there is any doubt as to whether a clinical observation is an AE, the event will be recorded. AEs and SAEs may also be identified via information from support departments e.g. laboratories or participant questionnaires.

The Symptoms questionnaire will be reviewed as soon as possible after completion and within 7 days if questionnaire is available and any clinically significant events identified by the PI (or delegated physician) will be recorded/reported as AEs/SAEs. A clinically significant event is defined as one which requires treatment and/or is recorded in the medical records.

Anaphylactoid reactions and cerebral oedema are adverse events of special interest (AESI) and will be recorded and reported. The DMC will review the occurrence of AESI and their related symptoms by treatment group at each meeting. Any cases of confirmed cerebral oedema confirmed by the Principal Investigator will immediately halt the trial and mandate DMC review before any further participants are randomised as this is a stopping criterion.

### Monitoring and oversight

The HiSNAP trial is an investigator-led study, sponsored by the Academic and Clinical Central Office for Research and Development for NHS Lothian/University of Edinburgh (ACCORD, reference AC23035). Trial oversight is directly provided by an independent Trial Steering Committee (TSC) and a separate independent DMC who will provide advice and review of safety.

### Study amendments and current status

The study protocol currently in effect is version 6 03 Jul 2024. A protocol version history is provided in supplement 1. Recruitment commenced 20 Mar 2024. At the time of authorship, 16 patients have been recruited and the trial is expected to complete recruitment on schedule.

### Ethics and dissemination

The trial will be conducted according to the ethical principles of the Declaration of Helsinki 2013 and was approved by East Midlands (Derby) Research Ethics Committee 26 Sep 2023 (reference 23/EM/0129), NHS Lothian Research and Development department, and the MHRA. Good Clinical Practice regulations will be followed and written informed consent will be obtained from all participants.

No trial results will be shared while the trial is ongoing. When the trial completes, data will be shared via presentation, publication and may be available on request from the trial Chief Investigator, decided on a case-by-case basis. Data sharing may be restricted in line with patent or commercial requirements for as short a time as possible.

Results will be disseminated by peer-reviewed publication, conferences, and linked on isrctn.com. Publication content will follow the Reporting of surrogate endpoints in randomised controlled trial reports (CONSORT-Surrogate) Extension of the CONSORT 2010 Statement.(19) Ownership of data arising from this study resides with the study team at the Sponsor Organisation. The study team will follow the International Committee of Medical Journal Editors guidelines on authorship. Requests for data access should be sent to the corresponding author.

## Supporting information

SPIRIT checklist

## Data Availability

Results will be disseminated by peer-reviewed publication, conferences, and linked on isrctn.com. Publication content will follow the Reporting of surrogate endpoints in randomised controlled trial reports (CONSORT-Surrogate) Extension of the CONSORT 2010 Statement. Ownership of data arising from this study resides with the study team at the Sponsor Organisation. The study team will follow the International Committee of Medical Journal Editors guidelines on authorship. Requests for data access should be sent to the corresponding author.

## Funding

The HiSNAP trial is an investigator-led study, funded by the Chief Scientist Office, part of the Scottish Government Health Directorates (reference TCS/21/04). For the purpose of open access, the author has applied a creative commons attribution (CC BY) licence to any author accepted manuscript version arising.

The investigators additionally acknowledge general expertise and support supplied pro bono by the Centre for Precision Cell Therapy for the Liver, a research centre funded by the Chief Scientist Office under the Precision Medicine Alliance for Scotland scheme.

### Authorship statement

All of the HiSNAP Trial Investigators authorship group made substantial contributions to the conception or design of the work; and either drafting the work or reviewing it critically for important intellectual content; and final approval of the version to be published; and all agree to be accountable for all aspects of the work in ensuring that questions related to the accuracy or integrity of any part of the work are appropriately investigated and resolved.

The HiSNAP Trial Investigators referenced in the authorship by-line are:

1. NHS Lothian, Royal Infirmary of Edinburgh, 51 Little France Crescent, Edinburgh, EH16 4SA, UK: Amy Armstrong, Caroline Blackstock, Oliver Carlill, Jonathan Carter, Donna Clark, Fraser Craig, Angela Crawford, Carlyn Davie, Craig Davidson, Judith Flemming, Julia Grahamslaw, Alison Grant, Alasdair Gray, Marie-Claire Harris, Katy Letham, Malgorzata Litwin, Fiona McCurrach, Shona McDonald, Fraser McFadyen, Hazel Milligan, Victoria Minnis, Lyle Moncur, Luisa Padovani, Gillian Pickering, Matt Reed, Rebecca Rielly, Emma-Beth Wilson, Philippa Wright, Lisa-Marie Butt.
2. Centre for Cardiovascular Science, University of Edinburgh, The Queen’s Medical Research Institute, 47 Little France Crescent, Edinburgh, EH16 4TJ, UK: Natalie Homer, Kathleen Scullion.
3. St. John’s Hospital, Howden W Rd, Howden, Livingston, EH54 6PP: Claire Cheyne, Angela McGeough, Nikki McLaughlin, Stephen Lynch, Fern Purdie.
4. Victoria Hospital, Hayfield Rd, Kirkcaldy, KY2 5AH: Sandee Beattie, Keith Boath, Anna Borkowska, Christina Coventry, Karen Gray, Chloe Haigh, Katie Hamilton, Jacqueline James, Keith Jacques, Jeronimo Martin Ramirez, Vasilika Ntoko, Surinder Pandhor, Sandra Pirie, Janine Ramsey, Heather Robertson, Maria Simpson, Alan Timmins

