## supplement 1 for "Study protocol: HiSNAP trial – a multi-centre, randomised, open label, blinded end-point, safety and efficacy trial of conventional (300mg/kg) versus higher doses of acetylcysteine (450mg/kg and 600mg/kg) in patients with paracetamol overdose in the United Kingdom"

| **Protocol version and implementation date** | **Changes** |
| --- | --- |
| Version 1 01 May 2023 | N/A |
| Version 2 28 Aug 2023 | - 5.6 - Information added - withdrawal of pt detained under Mental Health Act - 7.5 - Removal of storage of samples for future research - Other minor administrative changes |
| Version 3 12 Oct 2023 | - 5.3 – Definition of child-bearing potential added - 7.5 – Volume of urine samples updated - 11- Information added – review of symptoms Qs and reporting of AEs |
| Version 4 07 Nov 2023 | - 5.3 – Definition of post-menopausal updated. |
| Version 5 22 Mar 2024 | - Minor changes throughout to reflect modification from single to multicentre study. - Addition of ‘Summary of protocol changes’ table. - Other minor administrative changes |
| Version 6 03 July 2024 | - Substantial changes throughout to amend and add eligibility criteria, including changing from only acute paracetamol overdose to include patients with staggered or therapeutic excess, with presentation within 24 hours of taking last dose of paracetamol - Changes throughout to add mechanism to increase sample size beyond 90 patients - 2.2.2 - Removal of GLDH as secondary endpoint - 7.5 - Amendment from volume of blood tube to volume of blood taken and clarification of urine baseline sample to include pregnancy test sample - Other minor administrative changes |
